# Heterogeneity in network structure switches the dominant transmission mode of infectious diseases

**DOI:** 10.1101/2022.11.28.22282692

**Authors:** Pratyush K. Kollepara, Rebecca H. Chisholm, Joel C. Miller

**Author notes:** Designed research, performed research, analyzed data, wrote the paper (Pratyush K. Kollepara). Designed research, analyzed data (Rebecca H. Chisholm). Designed research, analyzed data; Address: PS-2, La Trobe University, Bundoora 3086, VIC, Australia |.

## Abstract

Several recent emerging diseases have exhibited both sexual and non-sexual transmission modes (Ebola, Zika and mpox). In the recent mpox outbreaks, transmission through sexual contacts appears to be the dominant mode of transmission. Motivated by this, we use an SIR-like model, to argue that an initially dominant sexual transmission mode can be overtaken by casual transmission at later stages, even if the basic casual reproduction number is less than one. Our results highlight the risk of intervention designs which are informed only by the early dynamics of the disease.

**Significance Statement:** The purpose of this article is to explore the risk from secondary transmission routes of diseases which spread through sexual contact. This is important because infectious diseases such as Ebola, Zika and mpox spread through both sexual transmission and other modes of transmission. Our results suggest that a secondary transmission route which is not dominant in the initial stages, can significantly alter the course of the epidemic and lead to more infections than expected in the later stages of the epidemic.

## I. INTRODUCTION

Mpox (also known as monkeypox) is a neglected tropical disease, endemic in Western and Central Africa, which frequently spills over from animal reservoirs into human populations. It has been documented to spread via respiratory droplets and skin-to-skin contact, which we refer to as ‘casual contact’ in this paper. The basic reproduction number for casual contact has been observed to be less than one [1–4]. In the recent outbreaks of this disease in non-endemic regions (mainly in the Americas and Europe), sexual contact was identified as the dominant cause of transmission with a reproduction number greater than one [5, 6].

A sexually transmitted disease where clearance of infection confers permanent immunity (in the absence of new susceptibles) would be expected to have a small final size compared to a non-sexually transmitted disease with the same basic reproduction number due to the highly heterogeneous contact structure of sexual transmission networks [7– 9]. However, if there is an additional transmission mechanism with a reproduction number less than one (ℛ < 1) [3, 4], we would expect that each individual infected through sexual contact would seed an outbreak through this additional mechanism which (on average) would haveℛ /(1 − ℛ) infections (subject to assuming the outbreaks are independent which would not be the case if the number of sexual transmissions makes up a sizeable proportion of the population). If ℛ is close to 1, the expected size of the outbreaks can be quite large. This raises concerns that an epidemic might be initially dominated by transmission through sexual contacts, but ultimately a large fraction (even the majority) of infections could occur through casual transmission (see Figure 1). In this paper, we explore the dynamics of such a disease, in which sexual transmission dominates at the start of the epidemic. We use a mathematical model to study the interplay between the transmission routes and analyze how their behavior changes.

**FIG. 1.**
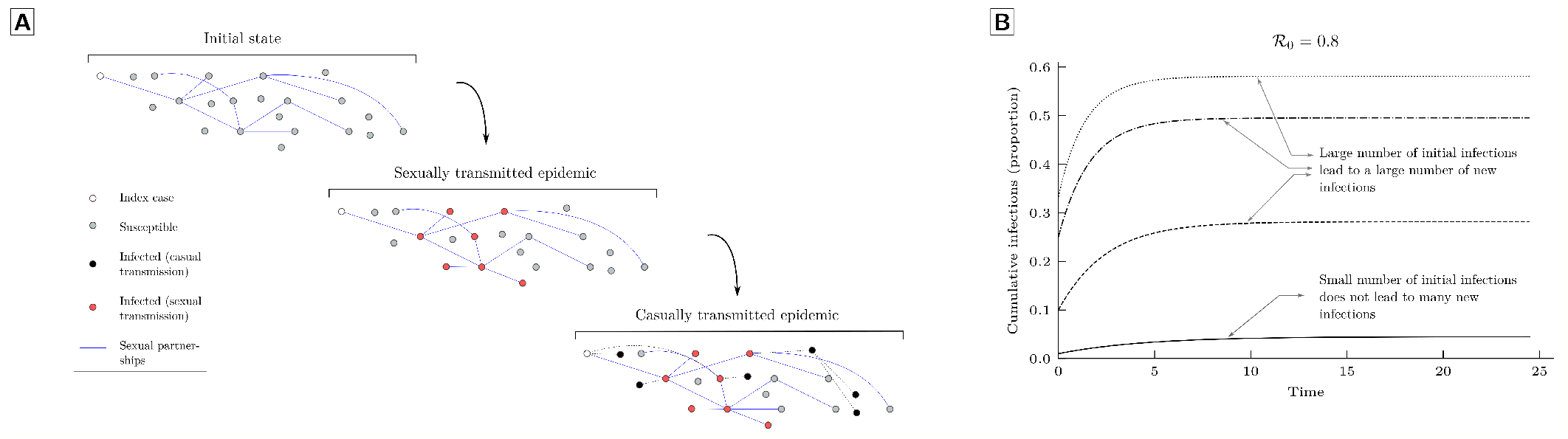
This figure illustrates the hypothesis that we explore in this paper. The ‘sexual’ epidemic (with a reproduction number greater than one) causes many infections in the population, each of which acts as the initial seed for the ‘casual’ epidemic (with reproduction number less than one), causing a significant number of infections. **(A)** The blue edges show the sexual partnership network that exists among the nodes. Casual transmission is homogeneous (equivalent a fully connected network), so the casual partnership network is not shown. Black dashed edges show casual transmission events. The initial state shows the population in which an infection is introduced (the index case). The ‘sexual epidemic’ state shows the disease transmitted along the sexual partnership network. The infections caused during the sexual epidemic act as a seeds for the ‘casual epidemic’ (for illustrative purposes we ignore sexual transmissions seeded from the ‘casual epidemic’). On average, an infected node leads to less than one direct casually transmitted infection, but the cumulative average number of descendants can be large. Further as there are many infected seeds from sexual transmission, the casually transmitted infections add up to a significant proportion of the population. **(B)** Time series of cumulative infections in a standard SIR model with ℛ_0_ = 0.8. This shows that if the initial number of infections is not small, a significant number of new infections can occur even if the reproduction number is less than one i.e. even if individuals on average cause less than one new infection.

Mpox is not the only disease with multiple transmission routes. Sexual transmission has been identified as a possible transmission route in Ebola and Zika in addition to their primary transmission route [10–15]. SARS-Cov-2 is airborne and can spread through contact with surfaces [16]. The neglected tropical disease, Chagas disease, is vector-borne and can spread through blood transfusions and oral routes [17, 18]. Scabies can spread via skin-to-skin contact and through fomites [19]. Trachoma can spread via close contact and is also vector borne [20].

This underlines the need for modeling of potential emerging diseases which have a casual contact reproduction number close to one and a sexual reproduction number greater than one. It would be expected that if the sexual transmission mode dominates at the start of the epidemic, intervention policies may focus only on sexual transmission and ignore transmissions from casual contacts. Despite the documentation of multiple transmission routes in several diseases, modeling literature on multiple transmission routes is sparse [21–23]. We adapt the framework developed in [23] to develop an SIR model that uses two routes of transmission: homogeneous mass action transmission (representing casual transmission) and a network with a heterogeneous degree distribution (representing sexual transmission).

## II. METHODS

### A. Compartmental Model

We build a model following an existing framework for sexual transmission and nonsexual transmission [23]. For sexual transmission, we use a heterogeneous mean-field (annealed) network. The annealed network assumption means an individual changes their contacts on a time scale that is faster than transmission, but maintains the number of contacts at a constant value. This assumption will later be used to derive the reproduction number in the next sub-section. For casual transmission, we assume that individuals mix homogeneously.

Starting with equations for the proportion of individuals in each disease stage, the model can be reduced to a system of four differential equations. We denote the proportion of the population with degree *k* (for the sexual network) in state *X* by *X*_*k*_, where *X* ∈ {*S, I, R* }. The proportion of the population in a state *X* (summed over all degrees) is denoted by *X*. The proportion of the population with degree *k* (summed over all states) is denoted as *N*_*k*_. The per-partnership sexual contact transmission rate is *β*_1_, the overall casual contact transmission rate is *β*_2_ and the recovery rate is *γ*. Our governing equations are

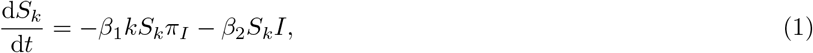

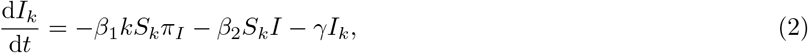

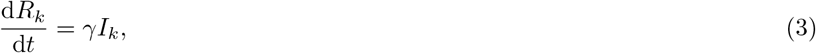

where 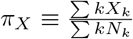 is the probability that a random sexual contact occurs with an individual in the state *X*. We can also define the probability generating function

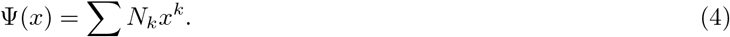

It can be immediately seen that Ψ^′^(1) = ∑ *kN*_*k*_ = ⟨*K*⟩ and Ψ^′′^(1) = ⟨*K*(*K* − 1)⟩. Using the differential equation for *S*_*k*_

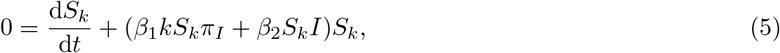

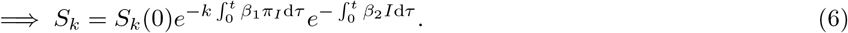

Let

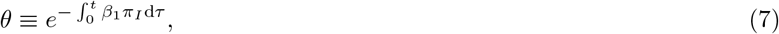

for which

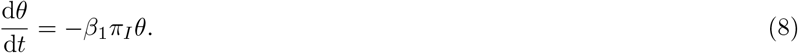

Similarly, let

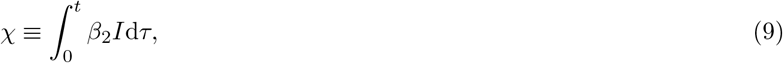

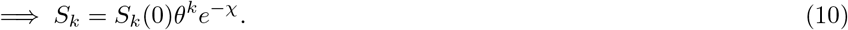

If we assume that only sexual transmission is present, then *θ*^*k*^ is the proportion of individuals of degree *k* that are susceptible. This proportion equals the probability that a randomly chosen individual of degree *k* is not infected, or in other words, was never exposed to transmission. Similarly, if only casual transmission is present then *e*^−*χ*^ is the probability of never being exposed to transmission. When both transmission modes are present, the probability that a randomly selected individual is not infected is equal to the product of the probability that the individual was never exposed to sexual transmission from an infected individual and the probability that the individual was never exposed to casual transmission from an infected individual. Therefore, *θ*^*k*^ and *e*^−*χ*^ can be interpreted as the probability of not being exposed to sexual transmission (for the given degree *k*) and casual transmission, respectively. For the initial conditions, we assume that the proportion of initial infections in the population is *ρ*_*k*_. We can now find an expression for *S*(*t*)

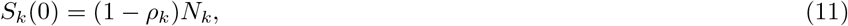

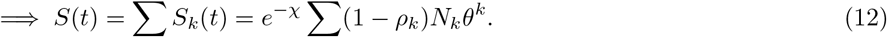

Similarly,

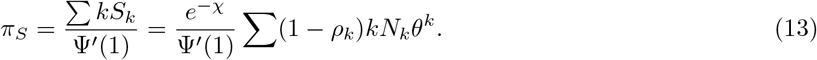

The system of equations (1, 2 and 3) can then be reduced to four differential equations

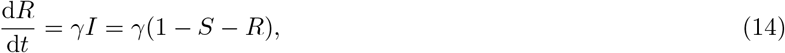

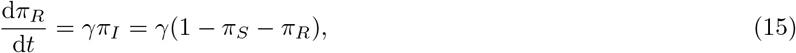

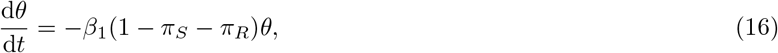

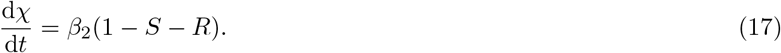

These equations can be solved numerically. In the limit *ρ*_*k*_ → 0, *S*(*t*) = Ψ(*θ*)*e*^−*χ*^. We can interpret Ψ(*θ*) as the probability of not having been exposed to sexual transmission, and *e*^−*χ*^ as the probability of not having been exposed to transmission through the casual contact route. In Figures 2, we plot 1 − Ψ(*θ*) and 1 − *e*^−*χ*^ as a function of time. These are the probabilities that a randomly chosen individual was exposed to sexual transmission at least once, or to casual transmission at least once, respectively. In solving the system of differential equations (14 - 17) numerically, we use *ρ*_*k*_ ∝ *k* as the initial conditions where all the *ρ*_*k*_ ≪ 1. This initial condition is used because we assume that an individual with more sexual contacts is more likely to get infected.

**FIG. 2.**
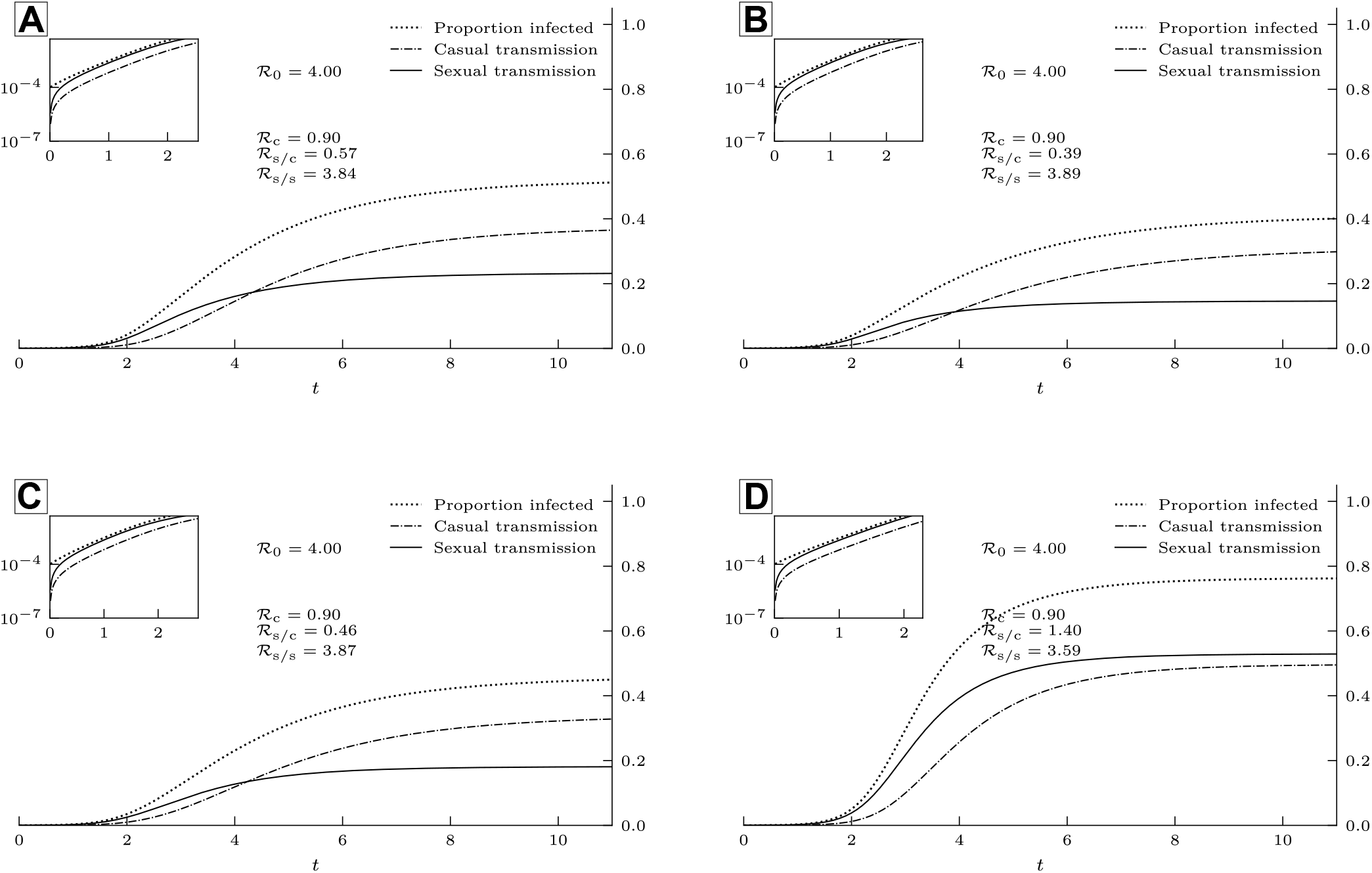
The probability of getting exposed to casual transmission may exceed that of sexual transmission even when the reproduction number for casual transmission is less than one (ℛ_c_ *<* 1 ℛ*<* _s/s_). Parameters: (**A**) *N*_0_ = 0.34, *k*_max_ = 60, *α* = 2; (**B**) *N*_0_ = 0.75, *k*_max_ = 20, *α* = 2; (**C**) *N*_0_ = 0.75, *k*_max_ = 60, *α* = 3; (**D**) *N*_0_ = 0.34, *k*_max_ = 20, *α* = 3. The recovery rate is *γ* = 1 for all cases. The reproduction numbers are as follows – ℛ_*c*_: casually transmitted infections caused by any individual, ℛ_s/s_: sexually transmitted infections caused by a sexually infected individual, ℛ_s/c_: sexually transmitted infections caused by a casually infected individual. We use ℛ_0_ = 4 and ℛ_*c*_ = 0.9 and the rest of the reproduction numbers are determined. The three curves show the probability that a randomly selected individual (i) is susceptible, given by 1 − *S* and shown by dotted line, (ii) was exposed to sexual transmission, given by 1 Ψ(*θ*) and shown by solid line, (iii) was exposed to casual transmission, given by 1 − *e*^−*χ*^ and shown by dot-dashed line. The inset is the initial dynamics at magnified scale with log-scale for the y-axis, which shows the dominance of sexual transmission over casual transmission. The cross over can be seen in (**A, B, C**) where casual transmission starts to dominate.

### B. Sexual transmission network

For the sexual transmission network, we use a class of networks whose degree distribution *P* (*K* = *k*) (we use *N*_*k*_ as a shorthand for this) is given by

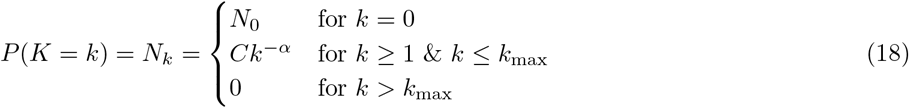

for integral values of *k*, where *N*_0_ is chosen arbitrarily, 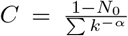 is a normalization constant and *k* _max_ is the maximum number of sexual contacts an individual is allowed to have. The exponent − *α* with *α >* 0 corresponds to decay at large *k*, but much slower than exponential. Often, scale-free networks or Erlang distributed networks are used for modeling of sexual transmission networks [24–26] to capture the slow decay at large *k*. Our network differs from these networks by including a cutoff on the degree and the presence of nodes with zero degree. In the next subsection, we show that the second moment of the distribution, ⟨ *K*^2^⟩, forms a component of ℛ_0_. The qualitative features of our results will persist irrespective of the details of the distribution. What is crucial is that the distribution has sufficient high-degree individuals that ⟨ *K*^2^ ⟩ is large compared to ⟨ *K* ^2^ ⟩, but the high-degree individuals comprise a small proportion of the population.

### C. Next generation matrix and ℛ_0_

The reproduction number can be obtained by calculating the largest eigenvalue of the next generation matrix *G* [27, 28], *whose elements are:*

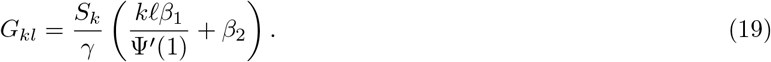

Another approach is by using a lower dimensional description of the system. At each generation, the newly infected population can be grouped into those who have been infected through sexual contact 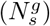 and those who have been infected through casual contact 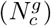. In the next generation, this grouping is transformed by a next generation matrix [29]

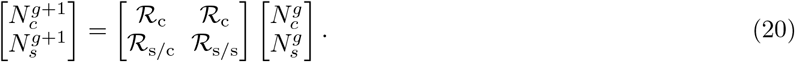

The average number of new infections caused by an individual through casual transmission is ℛ _c_. From the model, we see that the transmission rate is *β*_2_ for casual transmission and 1*/γ* is the time spent in the infectious stage, so 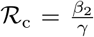. The average number of new sexually transmitted infections caused by an individual who was infected casually is 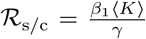. The average number of new sexually transmitted infections caused by an individual who was infected sexually is 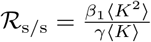. These can be computed by considering the following.

There are two network layers, a homogeneous layer and the sexual transmission layer with degree given by random variable *k* and a distribution *N*_*k*_. This leads to four types of reproduction numbers:

i. Casual infections caused by a casually infected individual: the casual contact network distribution is homogeneous with all transmitting at rate *β*_2_ with an average duration of 1*/γ*. So the expected number of new casual infections per infected individual is 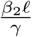.
ii. Casual infections caused by a sexually infected individual: the expected number of new casual infections is also 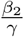, because being infected through sexual contact does not affect transmission through the contact network.
iii. Sexual infections caused by a sexually infected individual: The expected number of infections caused by a node of degree *k* is 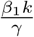 and the probability that a newly infected node has degree *k* is 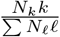 (higher the degree, higher is the chance of contacting an infected individual). Thus, the expected number of infections is proportional to 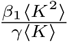.
iv. Sexual infections caused by a casually infected individual: the number of infections caused by a node of degree *k* is 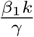. The probability that the newly infected node has degree *k* is *N*_*k*_, and the expected number of infections is 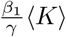.

Note that in (i) and (ii) above, the reproduction numbers are identical,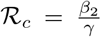.For (iii) and (iv), using the appropriate normalization, ℛ _s/s_ and ℛ _s/c_, are obtained respectively [29]. The largest eigenvalue of the matrix in equation (20) matches with that of the next generation matrix defined in equation (19).

### D. Final state relations

We can derive transcendental equations for the variables *θ* and *χ* at the end of the epidemic by assuming that the proportion of initial infections in the population is negligible, i.e., *ρ*_*k*_ → 0. Starting with equation (17) and using equation (14),

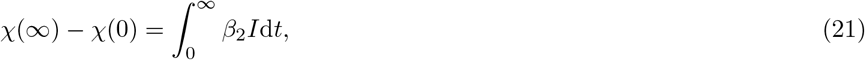

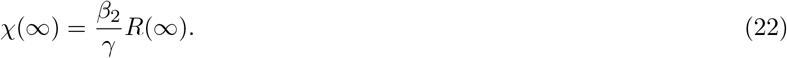

At the end of the epidemic, *I*(∞) = 0, and *S*(∞) can be obtained using equation (12)

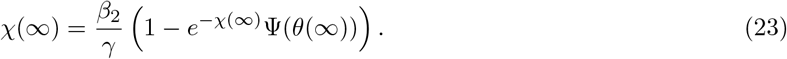

The equation for *θ* can be derived by considering its definition and using equation (15)

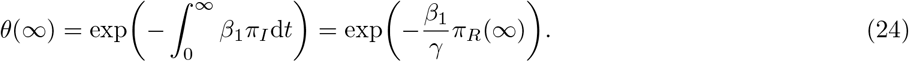

At the end of the epidemic, *π*_*I*_ (∞) = 0, and *π*_*S*_(∞) can be obtained from equation (13)

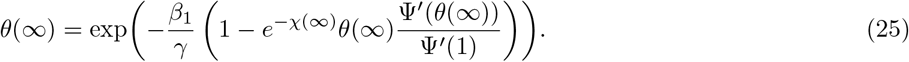

Thus, equations (23 and 25) together are transcendental equations for the final state of *χ* and *θ*. We solve them numerically through recursion and use the solutions for constructing Figure 3.

**FIG. 3.**
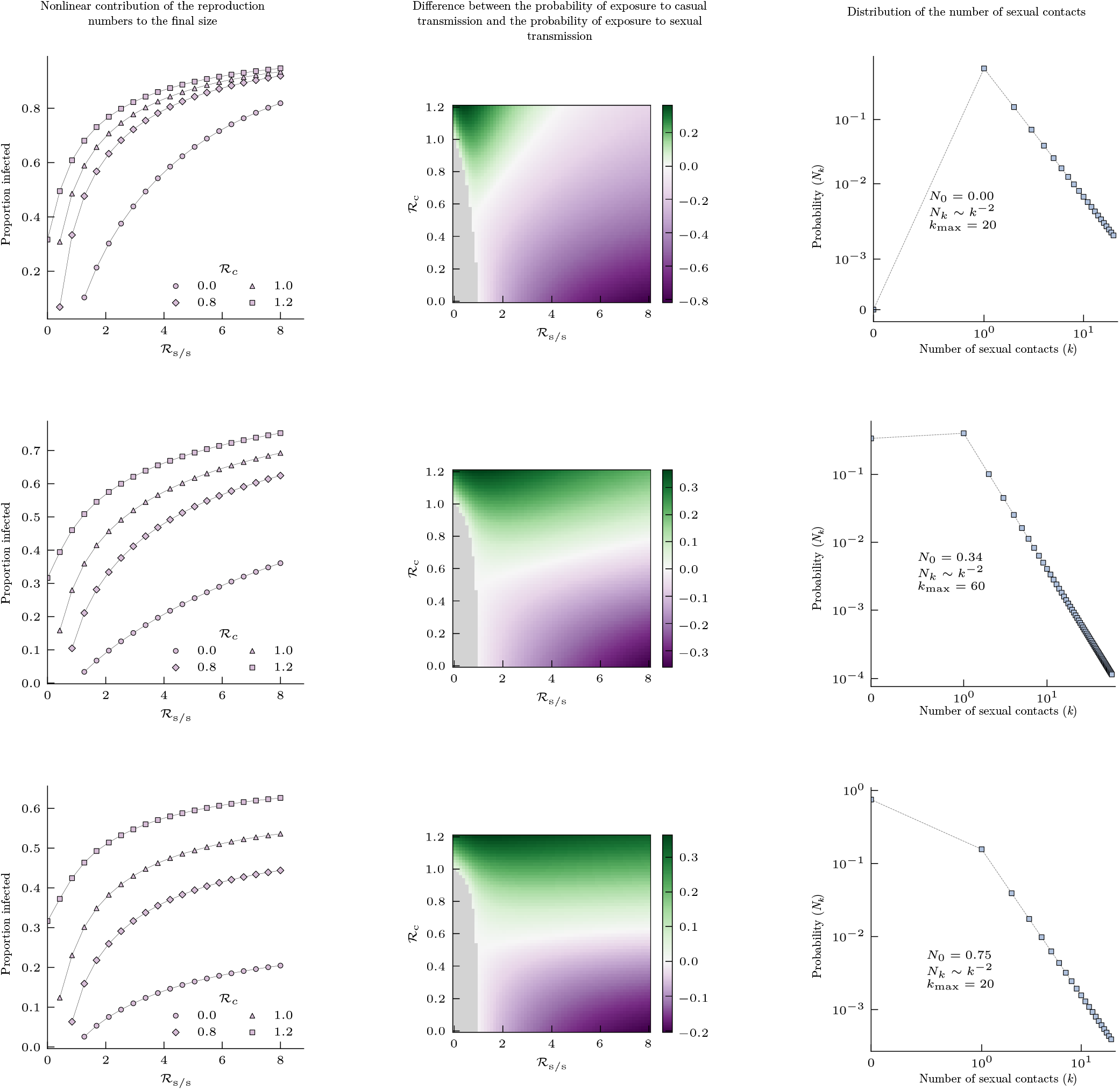
Sections of parameter space of the model. The first plot of each row shows how the final size or the proportion infected (*R*(∞)) in the epidemic, changes with ℛ_s/s_ for different values of ℛ_*c*_, the casual contact reproduction number. The second plot in each row shows a heat map of the difference between probability of exposure to casual transmission and probability of exposure to sexual transmissi, at the end of the epidemic, plotted across the basic casual reproduction number and basic sexualsexual reproduction number. Green regions in the heatmap indicate the reproduction numbers for which casual transmission eventually dominates. The third plot shows the degree distribution and its parameters. For all the three cases, the recovery rate *γ* = 1.

## III. RESULTS

We are interested in studying if the presence of casual transmission (through homogeneous mass action) with a basic reproduction number ℛ_*c*_ less than one, in addition to sexual transmission (through a heterogeneous network) with a basic reproduction number ℛ_s/s_ greater than one, can affect the dynamics of an epidemic. We use a heuristic method to find some long-term insights into this problem. An infectious individual would on average create _*c*_ infections, which would lead to 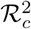 infections in the next generation, and so on, through casual contacts. For a large population, ignoring the fact that chains of transmission from multiple seeds may intersect and ignoring the sexual transmissions seeded by these chains, we can see that the total number of casually transmitted infections originating from an infected individual approaches the sum 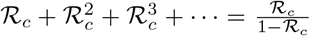. As ℛ_*c*_ → 1, the sum diverges and we would expect a large number of infections from casual contact for each ‘seed’ infected through sexual transmission. Casual transmission by a single infectious individual may not amount to a large proportion of infections, but when a significant proportion of the population is infected through sexual transmission, all the infections caused by casual transmission will add up to a considerable proportion. If ℛ_*c*_ *>* 1*/*2, then we can expect more than one casual infection per seed.

We study the same scenario using equations (14)–(17) that incorporates casual transmission using a mass action type dynamics and sexual transmission using an annealed network chosen to produce a power law-like degree distribution, described in the methods section. Our results show that the role played by the two transmission modes in the spreading of the disease at early times is not a good indicator of their roles at later times. The early dynamics are analyzed using the next generation matrix derived in equation (20). The stable distribution of transmissions, meaning the proportions of casual transmissions and sexual transmissions for each generation is obtained by the top eigenvector of the next generation matrix, 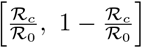. If ℛ_*c*_ ≪ ℛ_s/s_, then ℛ_0_ ≈ ℛ_s/s_ [23]. Therefore, more infections will be caused by sexual transmission in the early times.

In Figure 2 we show the trajectory of the epidemic for four populations that have different sexual contact networks with R_*c*_ = 0.9 and ℛ_0_ = 4. Since ℛ_*c*_ *<* ℛ_s/s_, the probability of getting exposed to sexual transmission (1 − Ψ(*θ*)) is initially higher than that of getting exposed to casual transmission (1 − *e*^−*χ*^). However, as the epidemic progresses, the dominant mode of transmission changes and the probability of exposure to casual transmission exceeds that of sexual transmission. This behavior, which we refer to as ‘switching of the dominant transmission mode’, can occur over a wide range of parameter values, and the final size can considerably change with the presence of casual transmission (Figure 3).

The mechanism for switching of the dominant transmission mode can be deduced from equation (12). The rate at which susceptibles deplete is higher for those with a larger degree. When the epidemic is starting, the first nodes to get infected sexually and recover are disproportionately those with the highest degree. Since the proportion of high degree nodes is very small (*N*_*k*_ ∼ *k*^−*α*^), after the initial stage, the epidemic is sustained only through the more prevalent low-degree nodes which do not contribute much to sexual transmission. Therefore, despite having a low basic reproduction number, the probability of exposure to casual transmission may be comparable to or even exceed the probability of exposure to sexual transmission in the later stages of the epidemic.

In contrast, if ℛ_*c*_ were larger than ℛ_s/s_, casual transmission would certainly dominate in the early times. However, as discussed above, sexual transmission events become less likely after a short period of time due to depletion of higher-degree susceptibles. So, casual transmission will continue to dominate throughout the epidemic. Thus, the heterogeneous degree distribution (of the initially dominant mode) is playing an important role in switching the dominance of transmission modes and just the existence of two routes of transmission with disparate reproduction numbers is not sufficient for switching.

Figure 3 also shows the disproportionate effect that casual transmission has on the final epidemic size. Despite the fact that ℛ_*c*_ is smaller than one, reducing it to zero may lead to a larger reduction in final size than reducing ℛ_s/s_ by the same amount, depending on the characteristics of the sexual contact network and values of the reproduction numbers.

## IV. DISCUSSION

In this paper, we have presented and analyzed a model of a disease with two transmission routes: casual and sexual, based on [23]. We model casual transmission using homogeneous mass action dynamics and sexual transmission using an annealed network with heterogeneous degree distribution. The degree distributions from empirically observed sexual transmission networks are often described as long-tailed distributions (*P* (*X* = *x*) ∼ *x*^−*α*^ with *X* > 0) [24, 25]. However, the social and biological constraints on an individual’s sexual activity would imply that there is an upper limit on the number of sexual contacts. To account for this, distributions which have an exponential cutoff for large degrees are often employed [24, 26]. Instead of an exponential cutoff, we impose a cutoff on the maximum degree and also allow for a non-zero probability for individuals with zero sexual contacts. We have approximated casual transmission using homogeneous mass action dynamics i.e. all individuals are identical. Casual transmission, although heterogeneous, has not been reported to have long-tail properties [30, 31]. Relative to the large dispersion in the distribution of sexual contacts, the dispersion in the casual contact distribution would be negligible, justifying the homogeneous assumption.

Sexual transmission as an additional route of transmission was identified for both Zika and Ebola viruses [10, 11, 13, 14]. Sexual transmission in mpox was a discovery from the recent outbreaks in non-endemic regions. From observations in endemic regions, it was believed to spread through casual contact and zoonotic transmissions and sexual contact was not documented as a transmission route [1, 2, 5, 6]. This work started out as an attempt to explore the potential impact of casual transmission in later stages of the emerging mpox epidemics, which were mainly driven by sexual contact transmission. But since the time of writing, the outbreaks have started to subside. At its peak in mid-August, about a thousand daily cases of mpox were detected world-wide. Heightened awareness of the disease, contact tracing, COVID-19-induced social distancing and a very small basic casual reproduction number may explain why the outbreaks did not continue to grow.

Although this analysis was originally motivated by mpox, our results are applicable to any SIR-like disease with a dominant sexual mode of transmission (basic reproduction number greater than one) and a secondary casual mode of transmission (basic reproduction number less than one). The main insight from our model is that those individuals with highest levels of sexual activity (and therefore most infectious) are at the highest risk of getting infected at the time of epidemic onset. But as the epidemic progresses, casual transmission can become dominant and even those who are sexually inactive can be at risk of getting infected.

In terms of designing intervention policies, recognizing the possibility of changing dominant transmission modes has important implications:

i. Expected outcome of interventions: Due to the heterogeneous degree distribution, the final size of a disease spreading on a sexual contact network is expected to be smaller than that in a homogeneous population, for the same value of ℛ_0_. Our results show that final size estimates based only on sexual transmission could be significant underestimates. Underestimating the final size in this manner could have a decisive impact on the performance of interventions. If the interventions are intensive and can completely eliminate sexual transmission at early stages of the epidemic, then the secondary route of casual transmission will not pose any significant risk. On the other hand, if the interventions are not able to eliminate, but only moderate sexual transmission, then casual transmission could become dominant at a later time leading to a significantly larger final size than expected. Thus, intervention policies for curtailing sexual transmission, based on the early observations of a disease may fail to meet the expected outcomes if an initially insignificant casual transmission route is ignored.
ii. Risk factors for different groups: For sexually transmitted diseases, children are generally considered to be not at-risk because they are sexually inactive. The degree distribution we have used accounts for a sexually inactive component of the population and the results show that this group is also at risk once casual transmission becomes dominant.
iii. A broader space of interventions: When ℛ_s/s_ ≫ ℛ_*c*_, sexual transmission forms the main component to the basic reproduction number and a reduction in ℛ_s/s_ will have an appreciable effect on reducing ℛ_0_. However, subject to the structure of sexual contacts, a reduction in ℛ_s/s_ and in ℛ_*c*_ by the same amount will have a starkly different effect on the final size of the epidemic. Even if the casual reproduction number is less than one, policymakers should consider the disproportionate contribution of casual transmission to the final size and implement interventions on casual transmission along with the interventions on sexual transmission.
iv. Critical community size: Diseases that induce immunity typically spread as an epidemic wave, and then infection counts crash for a long period until immunity wanes or new susceptibles join the at-risk population. In small populations the disease is likely to go extinct during this inter-epidemic phase [32]. The possibility of long chains of casual transmissions may help a disease to persist in smaller communities than would happen otherwise.

In conclusion, for a disease with dominant sexual transmission and a secondary casual transmission component at the beginning of the epidemic, the dominant transmission route can switch and by the end of the epidemic, casual transmission may have a far larger impact than what would be expected from only sexual transmission. Therefore, policymakers should consider interventions against casual transmission along with the conventional approaches to curtailing sexual transmission.

## Data Availability

All data produced in the present work are contained in the manuscript.
Simulation software for this modeling study are available at https://github.com/Joel-Miller-Lab/casual-sexual-transmission. Finalized versions of all software will be available on an external repository following final acceptance.

## ACKNOWLEDGMENTS

PKK was supported by La Trobe University Full Fee Research Scholarship and La Trobe University Graduate Research Scholarship. JCM was supported by startup funds provided by La Trobe University. Computer programs used in this paper are available at https://github.com/Joel-Miller-Lab/casual-sexual-transmission

